# Influence of Nurse’s Knowledge and Attitudes on Caring Behaviors for People with Opioid Use Disorder

**DOI:** 10.1101/2025.05.22.25328156

**Authors:** Inyene E. Essien-Aleksi, Yuan Zhang, Don Roosan, Tracie McPadden, Leslie Rideout, Michael Martin, Paula-Jo Beniers, Amy Lund, Danielle Leone-Sheehan, Alysse Wurcel

## Abstract

**Background:** Opioid use disorder (OUD) remains the leading cause of preventable substance use-related deaths in the U.S. Each year, more than 1 million Americans seek hospital care for OUD-related issues. However, they often receive inadequate treatment and face barriers such as limited overdose prevention education, delays in medication initiation, and insufficient post-discharge follow-up. Nurses are well-positioned to improve inpatient OUD treatment, yet qualitative research suggests they often have limited knowledge and negative attitudes toward OUD, which can decrease the level of compassion and empathy nurses have toward people with OUD. Validated instruments, such as the Care Behavior Index (CBI-6) and Drug and Drug User Problems Perceptions Questionnaires (DDPPQ), can be used to assess the relationship between nurses’ knowledge and attitudes and the level of care nurses provide people with OUD. This study examined the relationship between nurses’ knowledge/attitudes and caring. Additionally, we further examined differences in DDPPQ and CBI-6 scores by nurse’s demographic and work-related factors.

**Method:** A cross-sectional study was conducted from September 16^th^ to December 10^th^, 2024 at two Tufts Medicine hospitals in Eastern Massachusetts. Approximately 600 nurses are employed in the adult inpatient units of these hospitals. Nurses were recruited through emails, newsletters, and targeted in-person outreach to nursing leadership. The survey was collected online via REDcap. Participants demographics, work-related factors, knowledge and attitudes (DDPPQ), and caring behaviors (CBI-6) were measured. Simple linear and generalized linear model regression analyses were conducted to examine the relationship between DDPPQ and CBI-6 scores, adjusting for relevant covariates.

**Results:** Two hundred twenty-four nurses started the survey. A total of 125 nurses completed the entire survey and were included in the analysis (94% female, 82% white, 98% non-Hispanic, mean age 35 ± 11.69) years). Nurses’ perceived knowledge and attitudes were significantly associated with caring behaviors (β = –0.11, *p* < 0.0001), with higher DDPPQ scores (decreased knowledge and more negative attitudes) linked to reduced caring behaviors. Multiple workplace factors were associated with variations in nurses’ knowledge/attitudes and caring behaviors. However, holding an RN license was the only significant factor contributing to the relationship between knowledge/attitudes and caring.

**Conclusion:** Nurses’ knowledge and attitudes play a crucial role in shaping their caring behaviors toward people with OUD. Although multiple workplace factors were associated with nurses’ knowledge/attitudes and caring. The possession of an RN license was the only significant workplace factor contributing to the relationship between knowledge/attitude deficits and reduced caring. Given persistent gaps in hospital-based care and the growing prevalence of OUD, there is an urgent need for targeted interventions at both individual and organizational levels to improve nurses’ knowledge and attitudes and support the delivery of evidence-based compassionate care for people with OUD. Further research should explore how specific workplace factors influence nurse’s knowledge, attitudes, and training needs related to the care of people with OUD.

## INTRODUCTION

Medications for opioid use disorder (MOUD) have transformed hospital-based care for opioid use disorder (OUD), offering life-saving treatment that significantly reduces opioid-overdose deaths and OUD-related complications, including HIV and Hepatitis C infections, and criminal activity among people with OUD (7, 20, 21, 22, 25, 29). Each year, over 1 million adults in the U.S. seek hospital treatment for OUD (4). However, many people with OUD remain at heightened risk for post-hospitalization mortality due to limited access to overdose prevention education, delays in MOUD initiation, and inadequate post-discharge follow-up (4, 6). Although substantial progress has been made in developing and implementing hospital-based programs to improve post-hospitalization outcomes through interdisciplinary partnerships (4, 31), significant gaps in hospital care persist. Nurses are essential for developing sustainable public health interventions to address inequities in OUD care (4, 13). Currently, nurses comprise approximately 23% of the U.S. healthcare workforce and are uniquely positioned to implement evidence-based interventions at scale (3, 14, 20, 32). Despite the inpatient setting serving as a crucial access point for initiating evidence-based care for OUD, limited research has examined targeted interventions for nurses working in this environment. Previous research has shown that inadequate training, knowledge gaps, and personal biases can hinder nurses’ ability to deliver compassionate and evidence-based care to patients with substance use disorders (1, 2, 5, 15). Negative stigmatizing attitudes and misconceptions about evidence-based OUD care— particularly regarding MOUD use— remain a persistent barrier to effective nursing care across the United States (2, 15). An understudied factor is the relationship between nurses’ knowledge and attitudes toward people with OUD and their caring behaviors (36). Validated instruments, such as the Care Behavior Index (CBI-6) and Drug and Drug User Problems Perceptions Questionnaires (DDPPQ), can be used to assess the relationship between nurses’ knowledge and attitudes and the level of care nurses provide to people with OUD (9, 10, 34). The DDPPQ measures nurses’ self-perceived knowledge and attitudes toward people with drug-related problems (18, 33). The total DDPPQ score ranges from 20 to 140, with lower scores indicating more positive perceptions of knowledge and attitudes and higher scores reflecting more negative perceptions (33). The CBI-6 assesses the caring behaviors of nurses and other healthcare professionals (10, 12). Total CBI-6 scores range from 6 to 36, with higher scores indicating a greater perceived level of nurse caring (10). This study examined differences in nurses’ perceived knowledge and attitudes, and caring behaviors based on demographic and work-related variables, as well as the association between nurses’ knowledge and attitudes and their caring behaviors toward hospitalized people with OUD. Two research questions guided this study:

1. Are there differences in nurses’ perceived knowledge attitudes and caring behaviors toward people with OUD in adult inpatient settings based on their demographic and work-related characteristics?
2. What is the relationship between nurse’s perceived knowledge and attitudes and their caring behaviors toward people with OUD in adult inpatient settings?

## METHODS

### Study Design

We conducted a cross-sectional study to examine inpatient nurses’ self-reported knowledge and attitudes and their caring behaviors at two Tufts Medicine hospitals: non-profit community and teaching hospitals in Eastern Massachusetts. Eligible participants included nurses who self-reported to be aged 18 or older, working full-time, part-time, or per diem in adult inpatient units. Exclusion criteria included nurses not actively practicing at the bedside, travel nurses, nurse managers or administrators, and those who self-reported to have less than 6 months of inpatient experience. Convenience and purposive sampling methods were used to recruit participants who met the inclusion criteria. Institutional Review Board (IRB) approval from Merrimack College and Tufts Medicine was obtained before participant recruitment and data collection (IRB #00004941).

### Participants and Data Collection

Participants were recruited at Tufts Medicine between September 16 and December 10, 2024, through flyers posted in selected inpatient units, nursing grand rounds, newsletters, and meetings of the nurses’ leadership committee. Additionally, we utilized a HIPAA-compliant mobile application to recruit participants for the study. Surveys were distributed electronically via email using REDCap. Before participation, nurses were informed of the study’s purpose, procedures, risks, benefits, and incentives. The first page of the online survey included a written informed consent form, which participants were required to read and agree to before proceeding to the actual survey questions. No identifying information was collected. Participants were assured that their employer would not have access to their responses and that participation would not affect their employment status. The IRBs at Merrimack College and Tufts Medicine approved the study’s consent procedure. Upon survey completion, participants were entered into a raffle to win a $50 Amazon gift card. A total of 20 raffle prizes were offered.

### Study Measures

#### Dependent variable

*Caring Behaviors.* Nurses’ perception of caring behavior was measured using the revised Caring Behavior Inventory-6 (CBI-6) scale, originally developed by Wolf and colleagues (34, 35) and revised by Coulombe et al. (10). The CBI-6 assesses caring behaviors through six items, rated on a 6-point Likert scale ranging from 1 (“never”) to 6 (“always”). Participants were asked to rate their caring words and behaviors toward patients with OUD. Total scores range from 6 to 36, with higher scores indicating a greater perceived level of nurse caring. The CB1-6 has no specific cut-off points. The observed CBI-6 scores for the study cohort were between 21-36, with a mean score of 30.83 (± 4.05). Based on the observed CBI-6 range, we stratified CBI-6 scores into low (6–16), moderate (17–26), and high (27–36) levels. The CBI-6 has acceptable internal consistency ranging from Cronbach’s α = 0.89 to 0.89 (10, 12) and demonstrated good internal consistency reliability for this study sample (Cronbach α = 0.82).

#### Independent variable

*Knowledge and Attitudes*. Nurse’s perceived knowledge and attitudes toward people with OUD was assessed using the Drug and Drug User Problems Perceptions Questionnaires (DDPPQ). This instrument was originally developed by Cartwright (9) as the Alcohol and Alcohol Problems Perception Questionnaire (AAPPQ), a 30-item scale designed to measure healthcare professionals’ self-assessed knowledge and attitudes related to care provided to people with alcohol-related problems. The AAPPQ was later revised and validated by Watson et al. (33), renaming it as DDPPQ, to measure healthcare professionals’ self-assessed knowledge and attitudes toward care provided to people with drug-related problems. The DDPPQ consists of 20 items measuring the therapeutic attitudes and knowledge of healthcare providers using a 7-point Likert scale (1 = “strongly agree” and 7 = “strongly disagree”). The total DDPPQ score ranges from 20 to 140. Lower scores indicate more positive perceptions of health professional’s attitudes and knowledge, while higher scores indicate more negative perceptions (33). The DDPPQ includes five subscales: role adequacy, role support, role legitimacy, role-related self-efficacy, and job satisfaction. The DDPPQ has demonstrated good construct validity and internal consistency reliability in samples of nurses and other healthcare professionals (18, 33). Several studies have applied the total DDPPQ scores to measure the construct of: knowledge and attitudes. Prior research has used the DDPPQ in various ways: as total scores, subscale scores, or by categorizing items into knowledge and attitude dimensions (18, 23). However, DDPPQ does not have established cut-off points for interpretation. In this study, DDPPQ scores ranged between 21-99, with a mean score of 55.69 (SD ± 15.51). Based on this distribution, we stratified DDPPQ scores into four levels of perceived knowledge and attitudes: very low (81–99): Corresponding to Likert responses 4 (neither agree nor disagree) through 7 (strongly disagree), low (61–80): Likert responses 3 (agree) through 4 (neither agree nor disagree), moderate (41–60): Likert responses 2 (somewhat agree) through 3 (agree) and high (21–40): Likert responses 1 (strongly agree) through 2 (somewhat agree). The DDPPQ demonstrated high internal consistency in this study sample (Cronbach’s α = 0.88).

#### Covariates

We chose covariates based on their hypothesized association with the outcome variables. Demographic variables included age (dichotomized into age groups), gender, race (White, Black, Asian, or other), and self-reported ethnicity (Hispanic vs. non-Hispanic). Work-related variables included type of nursing license (RN, LPN, or nurse educator), employment status (full-time, part-time, or per diem), educational level (some college, associate degree, bachelor’s degree, or master’s degree), years of work experience (less than 1 year, 1–5 years, 6– 10 years, or more than 15 years), shift schedule (permanent day, permanent evening, permanent night, and rotating shift), type of hospital (university/teaching vs. community), and how often respondent cared for patients with OUD (past work-related OUD exposure). Covariates significantly associated with the outcome variable were adjusted in the regression analysis.

### Data Analysis

Data analyses were conducted using SAS version 9.4 (SAS Institute, Cary, NC). Composite scores for DDPPQ and CBI-6 were first calculated. Descriptive analyses were conducted for socio-demographic and work-related variables, with continuous variables reported as mean ± standard deviation (SD) and categorical variables expressed as percentages (%). Bivariate analyses were conducted using one-way ANOVA to examine differences in DDPPQ and CBI-6 scores across categorical variables with three or more groups. For demographic and work-related variables that showed significant differences in DDPPQ and CB1-6 scores, post hoc comparisons were conducted using the Tukey-Kramer test to identify specific sub-group differences. Two-sided significance was reported at *p* ≤ 0.05. An independent *t*-test was conducted to examine differences in DDPPQ and CBI-6 scores across demographic and work-related variables at two levels. Simple Linear Regression (Model 1) and Generalized Linear Models (Model 2) regression analyses were conducted to assess whether perceived knowledge and attitudes were associated with caring behaviors, adjusting for covariates found to be significant in the bivariate analyses.

## RESULTS

### Descriptive and Bivariate analyses

Two hundred twenty-four people started the surveys. A total of 125 participants completed the entire survey and were included in this analysis. Details about the study participants can be seen in Table 1. The mean age of participants was 35 years (SD ±11.69). The majority of the sample were female (94%), white (82%), and full-time employees (87%), with 41% working rotating shifts. Most participants held an RN license (98%), and 38% reported caring for people with OUD at least once a week.

**Table 1.**
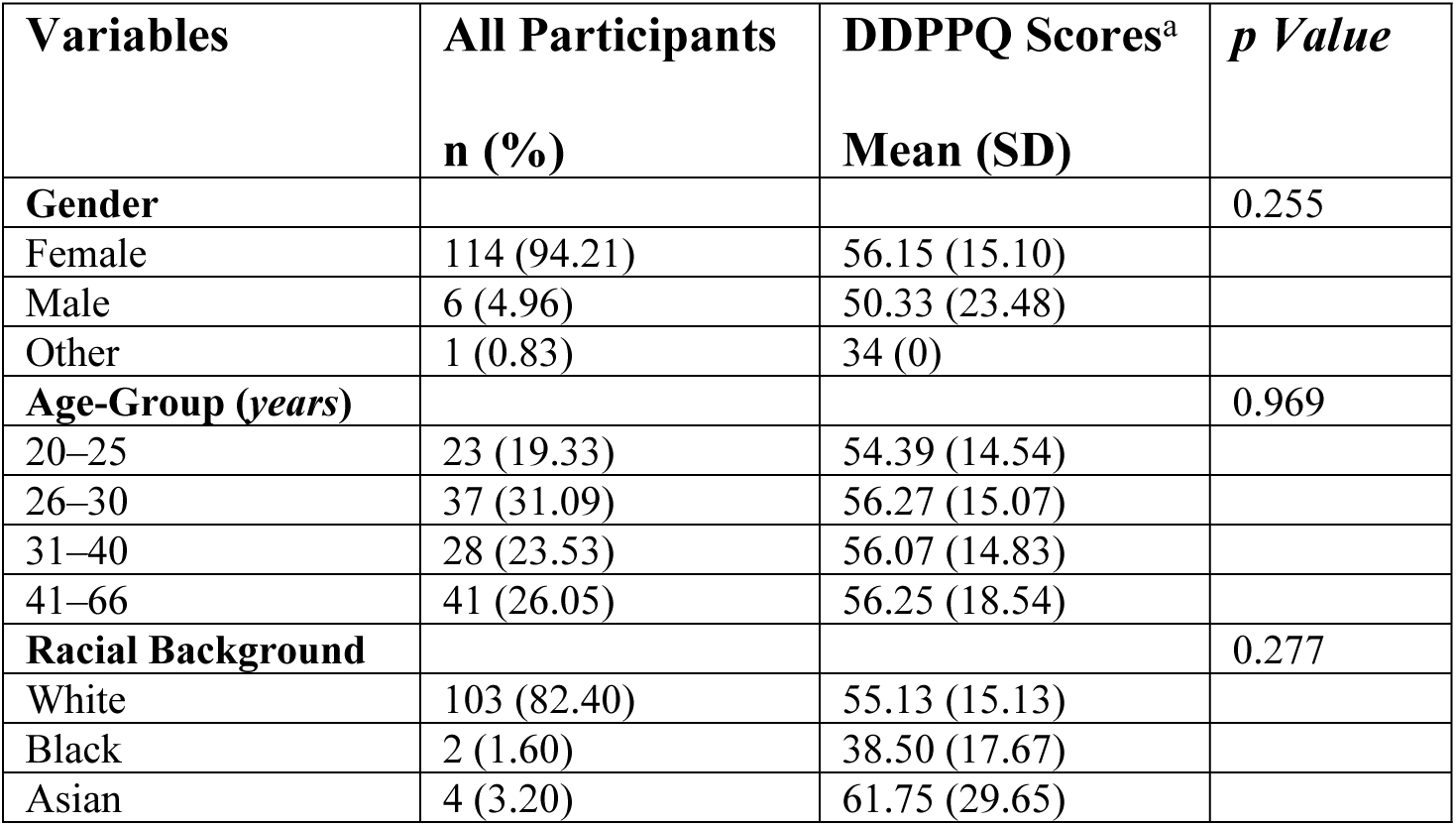
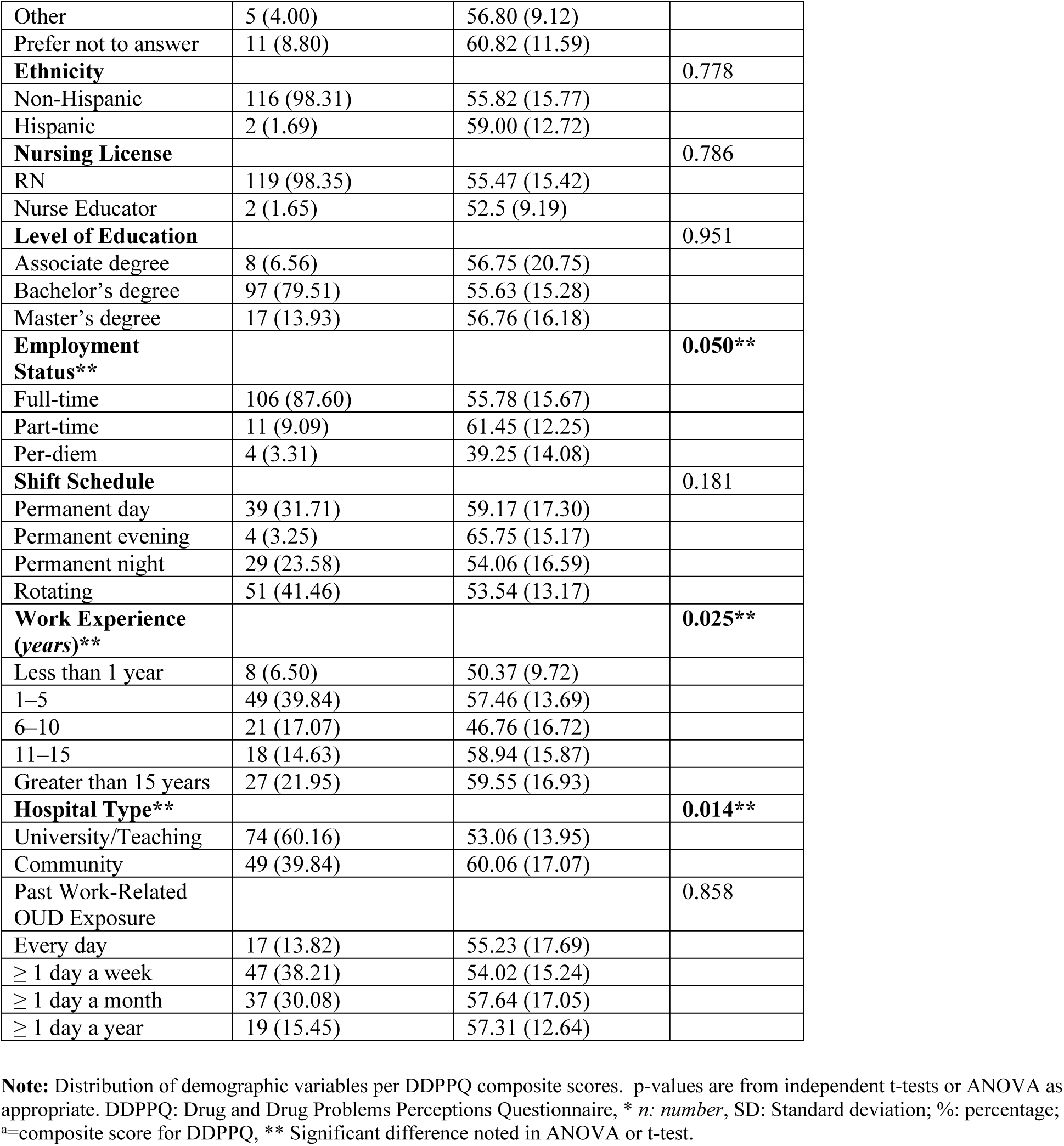
Participant Characteristics and Differences in DDPPQ Scores Across Demographic and Work-Related Variables (*n*=125)

| **Variables** | **All Participants**<br>**n (%)** | **DDPPQ Scores<sup>a</sup>**<br>**Mean (SD)** | ***p Value*** |
| --- | --- | --- | --- |
| **Gender** |  |  | 0.255 |
| Female | 114 (94.21) | 56.15 (15.10) |  |
| Male | 6 (4.96) | 50.33 (23.48) |  |
| Other | 1 (0.83) | 34 (0) |  |
| **Age-Group (years)** |  |  | 0.969 |
| 20–25 | 23 (19.33) | 54.39 (14.54) |  |
| 26–30 | 37 (31.09) | 56.27 (15.07) |  |
| 31–40 | 28 (23.53) | 56.07 (14.83) |  |
| 41–66 | 41 (26.05) | 56.25 (18.54) |  |
| **Racial Background** |  |  | 0.277 |
| White | 103 (82.40) | 55.13 (15.13) |  |
| Black | 2 (1.60) | 38.50 (17.67) |  |
| Asian | 4 (3.20) | 61.75 (29.65) |  |
| Other | 5 (4.00) | 56.80 (9.12) |  |
| Prefer not to answer | 11 (8.80) | 60.82 (11.59) |  |
| **Ethnicity** |  |  | 0.778 |
| Non-Hispanic | 116 (98.31) | 55.82 (15.77) |  |
| Hispanic | 2 (1.69) | 59.00 (12.72) |  |
| **Nursing License** |  |  | 0.786 |
| RN | 119 (98.35) | 55.47 (15.42) |  |
| Nurse Educator | 2 (1.65) | 52.5 (9.19) |  |
| **Level of Education** |  |  | 0.951 |
| Associate degree | 8 (6.56) | 56.75 (20.75) |  |
| Bachelor's degree | 97 (79.51) | 55.63 (15.28) |  |
| Master's degree | 17 (13.93) | 56.76 (16.18) |  |
| **Employment Status\*\*** |  |  | **0.050\*\*** |
| Full-time | 106 (87.60) | 55.78 (15.67) |  |
| Part-time | 11 (9.09) | 61.45 (12.25) |  |
| Per-diem | 4 (3.31) | 39.25 (14.08) |  |
| **Shift Schedule** |  |  | 0.181 |
| Permanent day | 39 (31.71) | 59.17 (17.30) |  |
| Permanent evening | 4 (3.25) | 65.75 (15.17) |  |
| Permanent night | 29 (23.58) | 54.06 (16.59) |  |
| Rotating | 51 (41.46) | 53.54 (13.17) |  |
| **Work Experience (years)\*\*** |  |  | **0.025\*\*** |
| Less than 1 year | 8 (6.50) | 50.37 (9.72) |  |
| 1–5 | 49 (39.84) | 57.46 (13.69) |  |
| 6–10 | 21 (17.07) | 46.76 (16.72) |  |
| 11–15 | 18 (14.63) | 58.94 (15.87) |  |
| Greater than 15 years | 27 (21.95) | 59.55 (16.93) |  |
| **Hospital Type\*\*** |  |  | **0.014\*\*** |
| University/Teaching | 74 (60.16) | 53.06 (13.95) |  |
| Community | 49 (39.84) | 60.06 (17.07) |  |
| Past Work-Related OUD Exposure |  |  | 0.858 |
| Every day | 17 (13.82) | 55.23 (17.69) |  |
| ≥ 1 day a week | 47 (38.21) | 54.02 (15.24) |  |
| ≥ 1 day a month | 37 (30.08) | 57.64 (17.05) |  |
| ≥ 1 day a year | 19 (15.45) | 57.31 (12.64) |  |

**Note:** Distribution of demographic variables per DDPPQ composite scores. p-values are from independent t-tests or ANOVA as appropriate. DDPPQ: Drug and Drug Problems Perceptions Questionnaire, \* *n*: number, SD: Standard deviation; %: percentage; <sup>a</sup>=composite score for DDPPQ, \*\* Significant difference noted in ANOVA or t-test.

*DDPPQ score:* The mean DDPPQ score was 55.69 (SD = 15.51), with scores ranging from 21 to 99. DDPPQ scores were stratified into four levels: very low (81–99), low (61–80), moderate (41–60), and high (21–40). Analyses using one-way ANOVA and independent *t*-test revealed no statistically significant differences in DDPPQ scores by age, gender, race, ethnicity, past work-related OUD exposure, type of nursing license, shift schedule, or educational level. Table 2 presents significant subgroup differences identified through pairwise comparisons using the Tukey-Kramer test (for ANOVA) and independent t-tests for DDPPQ scores across demographic and work-related variables. Nurses working at the university/teaching hospital had significantly lower DDPPQ scores than those at the community hospital (mean difference = –6.99, 95% CI [– 12.56 to –1.42], *p* = 0.014). This finding indicates that nurses in the university/teaching hospital reported higher perceived knowledge and more positive attitudes toward OUD care compared to their community hospital counterparts. We also found a significant difference between part-time vs. per-diem nurses, with part-time nurses scoring higher by 22.2 points (95% CI [ 0.89 to 43.51], *p* < 0.05), suggesting lower knowledge and more negative attitudes among part-time nurses compared to per-diem counterparts. In terms of work experience, nurses with 6-10 years of experience scored significantly lower than those with more than 15 years of experience (mean difference = –12.79, 95% CI: [-24.98 to –5.99], *p* = 0.025), indicating that more experienced nurses (those with more than 15 years of experience) did not report greater knowledge or more favorable attitudes toward people with OUD compared to mid-level experienced counterparts.

**Table 2.** Pairwise Comparisons of DDPPQ Scores by Hospital Type, Employment Status, and Work Experience.

| Comparison Variables | Groups compared | Mean Difference (95% CI) | *p* Value |
| --- | --- | --- | --- |
| **Hospital Type** | University/Teaching vs. Community | -6.99 (-12.56, -1.42) | **0.014\*** |
| **Employment Status** | Part-time vs. Per-diem | 22.20 (0.89, 43.51) | **0.050\*** |
| **Work Experience (years)** | 6–10 years vs. Greater than 15 years | -12.79 (-24.98, -5.99) | **0.025\*** |

**Note:** Mean differences were obtained using the Tukey-Kramer test (for ANOVA) and independent t-tests. DDPPQ = Drug and Drug Problems Perceptions Questionnaire. CI = Confidence Interval. Bold values are significant at the 95% CI level. \* Indicates statistically significant result at the $p < 0.05$ .

*CBI scores:* Table 3 presents significant subgroup differences identified through pairwise comparisons using the Tukey-Kramer test (for ANOVA) and independent t-tests for CB1-scores across demographic and work-related variables. The mean CB1-6 score was 30.83 (SD = 4.05), with scores ranging from 9-30. CB1-6 scores were categorized into three levels: low (9–16), moderate (17–23), and high (24–30) caring behavior levels. Analyses using one-way ANOVA and independent t-tests revealed no statistically significant differences in CB1-6 scores by age group, gender, hospital type, racial background, ethnicity, education level, past work-related OUD exposure, and employment status. One-way ANOVA only revealed significant differences in CBI-6 scores by shift schedule and type of nursing license; however, post hoc analyses showed no significant pairwise differences among specific shift types. In contrast, significant differences were observed based on the type of nursing license, with registered nurses (RNs) reporting higher CBI-6 scores than nurse educators (mean difference = 6.41, 95% CI [0.90, 11.92], *p* = 0.023). This finding indicates that RNs primarily working at the bedside displayed stronger caring behaviors than those in nursing education.

**Table 3.**
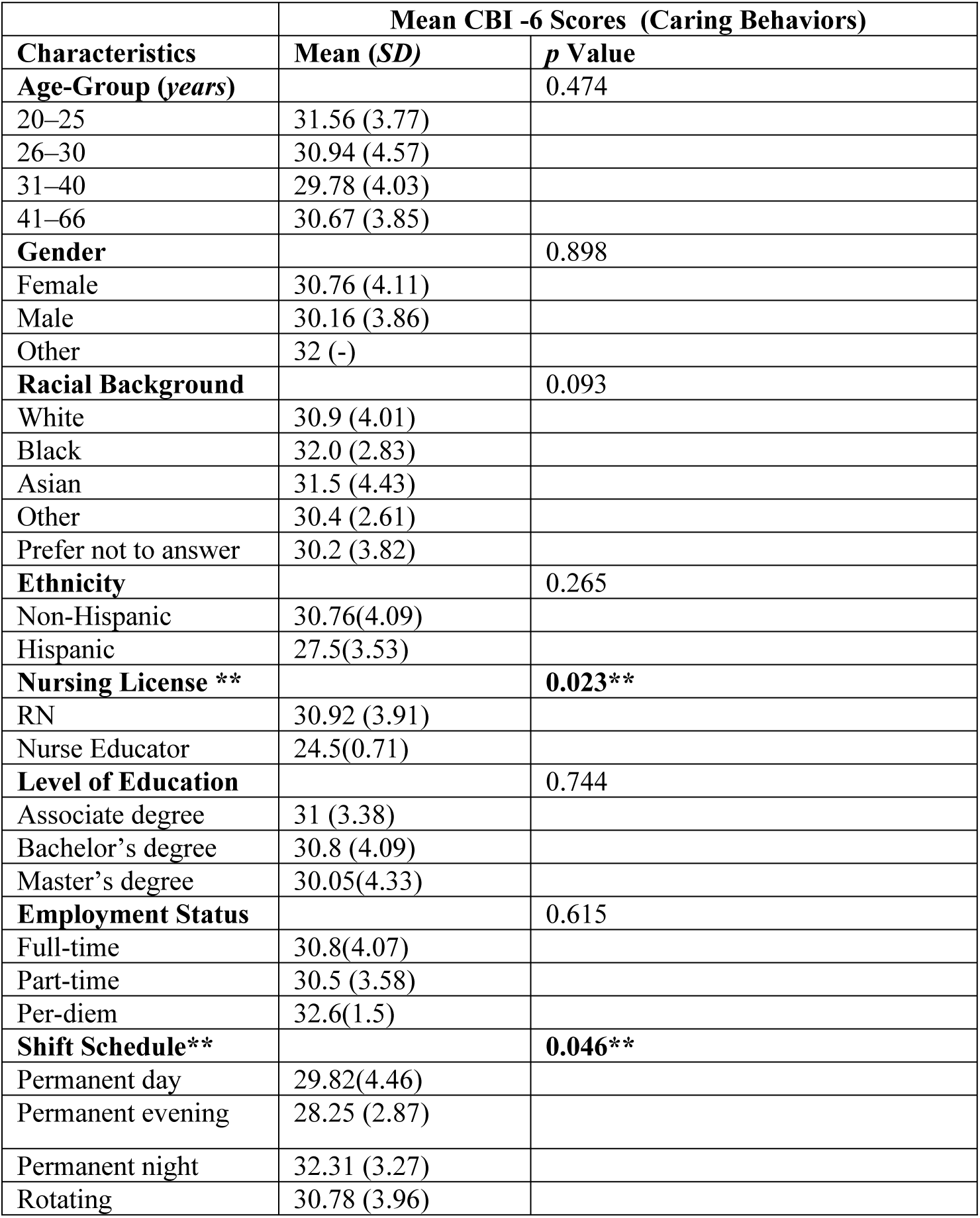
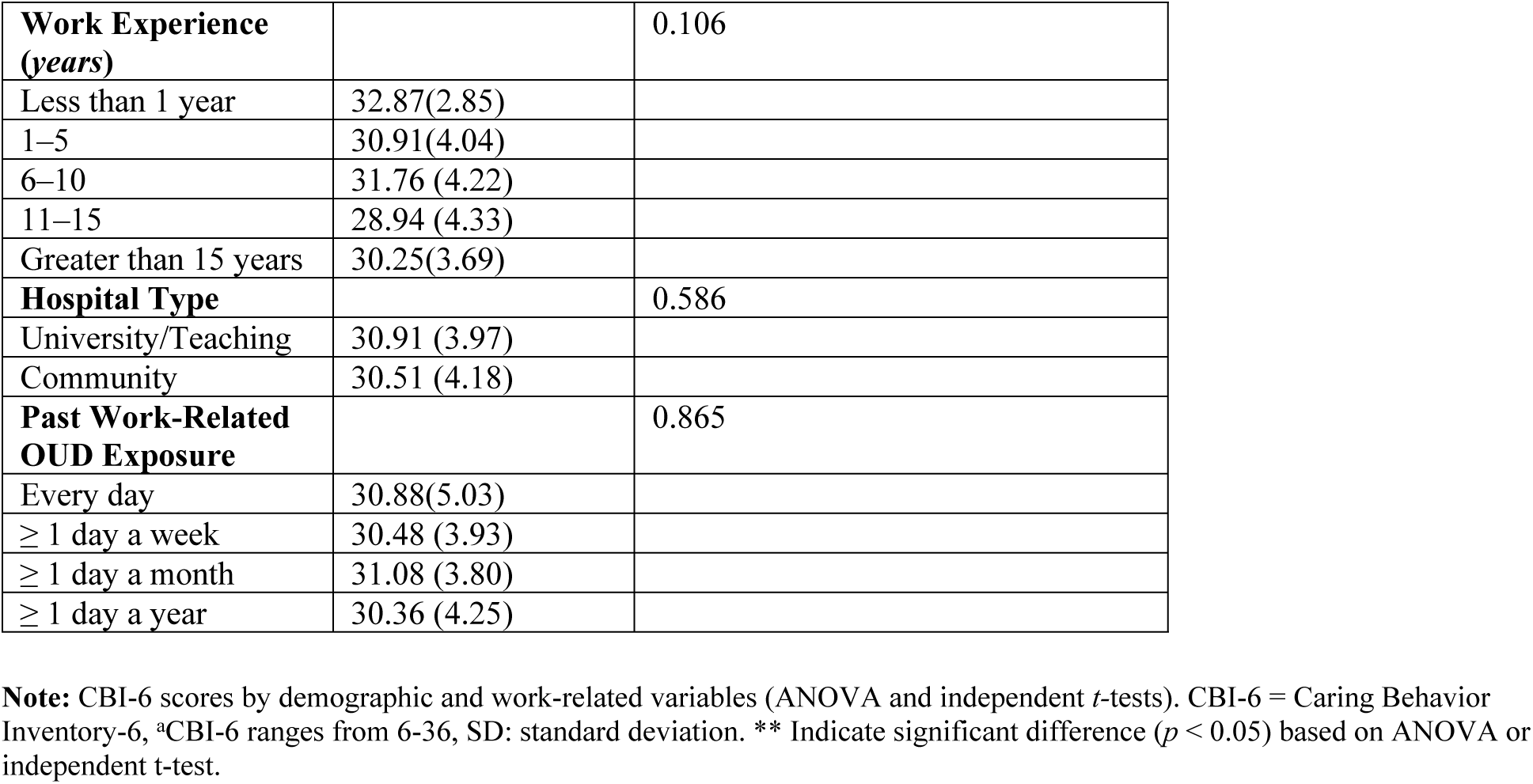
Differences in CB1-6 Scores Across Demographic and Work-Related Variables (*n*=125)

| **Characteristics** | **Mean CBI -6 Scores (Caring Behaviors)** |  |
| --- | --- | --- |
|  | **Mean (SD)** | ***p* Value** |
| **Age-Group (years)** |  | 0.474 |
| 20–25 | 31.56 (3.77) |  |
| 26–30 | 30.94 (4.57) |  |
| 31–40 | 29.78 (4.03) |  |
| 41–66 | 30.67 (3.85) |  |
| **Gender** |  | 0.898 |
| Female | 30.76 (4.11) |  |
| Male | 30.16 (3.86) |  |
| Other | 32 (-) |  |
| **Racial Background** |  | 0.093 |
| White | 30.9 (4.01) |  |
| Black | 32.0 (2.83) |  |
| Asian | 31.5 (4.43) |  |
| Other | 30.4 (2.61) |  |
| Prefer not to answer | 30.2 (3.82) |  |
| **Ethnicity** |  | 0.265 |
| Non-Hispanic | 30.76(4.09) |  |
| Hispanic | 27.5(3.53) |  |
| **Nursing License \*\*** |  | **0.023\*\*** |
| RN | 30.92 (3.91) |  |
| Nurse Educator | 24.5(0.71) |  |
| **Level of Education** |  | 0.744 |
| Associate degree | 31 (3.38) |  |
| Bachelor's degree | 30.8 (4.09) |  |
| Master's degree | 30.05(4.33) |  |
| **Employment Status** |  | 0.615 |
| Full-time | 30.8(4.07) |  |
| Part-time | 30.5 (3.58) |  |
| Per-diem | 32.6(1.5) |  |
| **Shift Schedule\*\*** |  | **0.046\*\*** |
| Permanent day | 29.82(4.46) |  |
| Permanent evening | 28.25 (2.87) |  |
| Permanent night | 32.31 (3.27) |  |
| Rotating | 30.78 (3.96) |  |
| **Work Experience (years)** |  | 0.106 |
| Less than 1 year | 32.87(2.85) |  |
| 1–5 | 30.91(4.04) |  |
| 6–10 | 31.76 (4.22) |  |
| 11–15 | 28.94 (4.33) |  |
| Greater than 15 years | 30.25(3.69) |  |
| **Hospital Type** |  | 0.586 |
| University/Teaching | 30.91 (3.97) |  |
| Community | 30.51 (4.18) |  |
| **Past Work-Related OUD Exposure** |  | 0.865 |
| Every day | 30.88(5.03) |  |
| ≥ 1 day a week | 30.48 (3.93) |  |
| ≥ 1 day a month | 31.08 (3.80) |  |
| ≥ 1 day a year | 30.36 (4.25) |  |

**Note:** CBI-6 scores by demographic and work-related variables (ANOVA and independent *t*-tests). CBI-6 = Caring Behavior Inventory-6, \*CBI-6 ranges from 6-36, SD: standard deviation. \*\* Indicate significant difference ( $p < 0.05$ ) based on ANOVA or independent *t*-test.

**Table 4.**
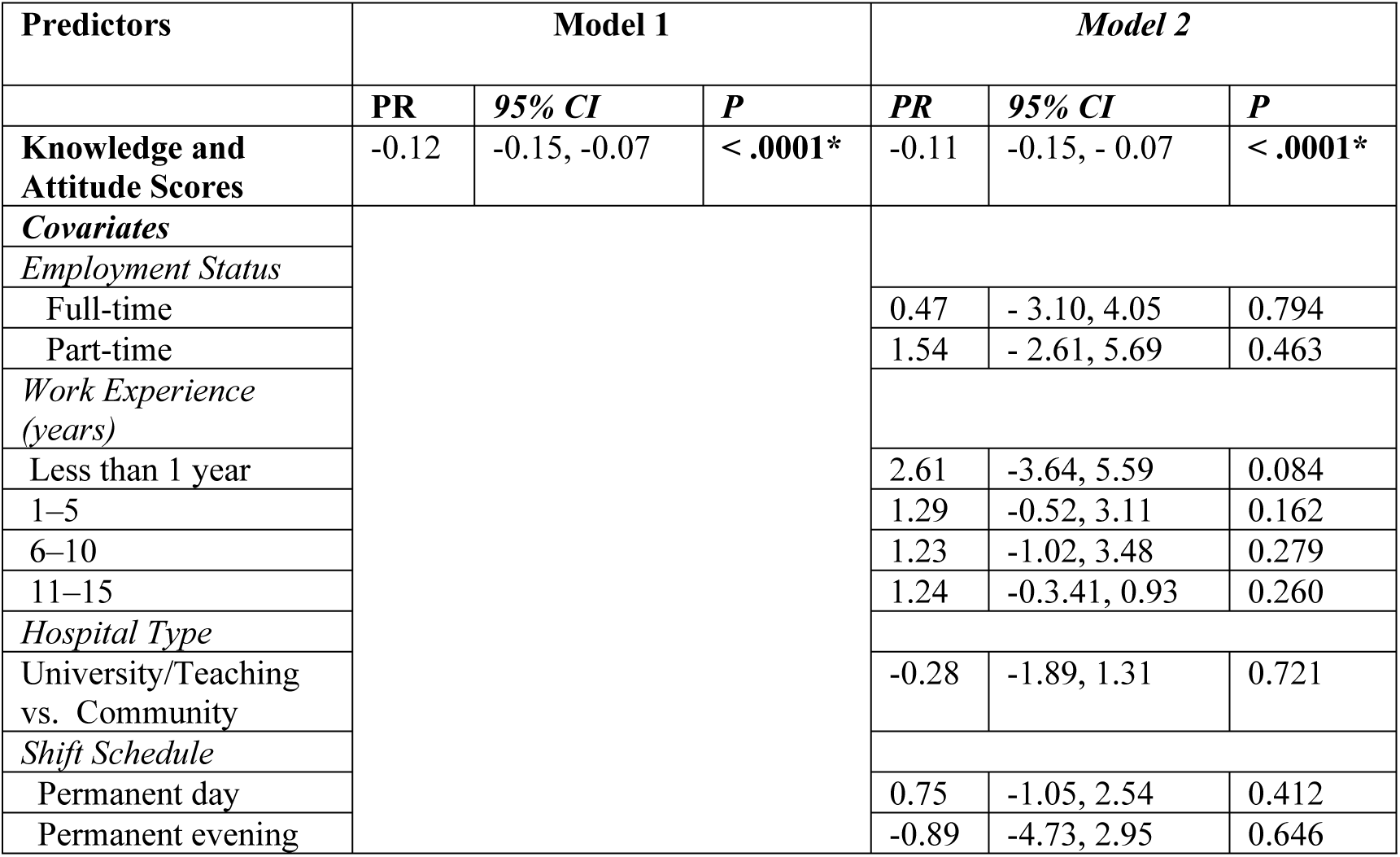
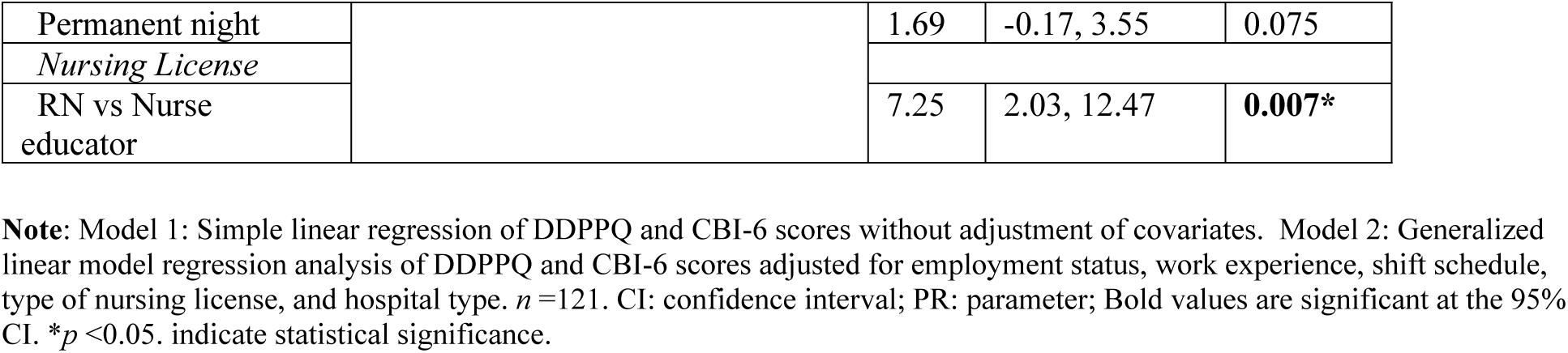
Regression Analyses of Nurse’s Knowledge/Attitudes and Work-Related Factors Predicting Caring Behaviors.

### Relationship Between Nurses’ Knowledge and Attitudes and Caring Behaviors

A simple linear regression analysis was conducted to examine whether nurses’ perceived knowledge and attitudes were associated with their caring behaviors toward people with OUD without adjustment (Model 1). The overall model was statistically significant, F(1, 123) = 30.55, *p* < .0001, indicating that DDPPQ scores significantly predicated caring behaviors. The regression model accounted for approximately 20% of the variance in respondents’ caring behavior scores (R² = 0.199, adjusted R² = 0.192). Nurses’ knowledge and attitude scores were negatively associated with their caring behaviors (β = –0.12, *p* < .0001), indicating that higher DDPPQ scores (lower knowledge and more negative attitudes) were associated with lower caring behaviors. A Generalized Linear Model regression analysis was conducted to further examine the relationship between nurses’ knowledge and attitudes and caring behaviors, adjusting for demographic and work-related covariates significantly associated with DDPPQ and CBI-6 scores in bivariate analyses (Model 2). These covariates included hospital type, employment status, work experience, shift schedule, and the type of nursing license. The regression model remained statistically significant, F(12, 107) = 4.56, *p* < .0001, accounting for 34% of the variance in caring behavior scores (R² = 0.34, adjusted R² = 0.29). Among the predictors, DDPPQ remained the strongest predictor of caring behaviors (β = –0.11, *p* < 0.0001), indicating that higher DDPPQ scores (poorer attitudes and less knowledge) were associated with lower caring behaviors toward people with OUD. Covariates, including hospital type, employment status, work experience, and shift schedule, were not significant in the model. The type of nursing license was the only significant predictor. Specifically, nurses holding only an RN license reported significantly higher CBI-6 scores compared to nurse educators (β = 7.25, *p* = 0.007). Overall, our study indicates that nurses’ knowledge and attitudes were the strongest predictors of their caring behaviors. Workplace factors, specifically the possession of an RN license, contributed to this relationship; whereas, hospital type, employment status, work experience, and shift schedule did not significantly influence nurses’ caring behaviors.

### Discussion

We found that higher scores on the DDPPQ—signals of lower knowledge and more negative attitudes—were significantly associated with reduced nursing caring behaviors toward people with OUD. This association remained robust and statistically significant after adjusting for demographic and work-related factors. While the possession of an RN license contributed to this relationship, other work-related variables such as hospital type, employment status, years of experience, and shift schedule, did not significantly influence caring behaviors. These findings contribute to the growing body of literature emphasizing the role of nurses’ knowledge and attitudes in shaping the quality of care provided to people with OUD. Although previous research has demonstrated that nurses often hold negative attitudes and limited knowledge about individuals with substance use disorders (1, 18, 36), the behavioral implications of these deficits have been largely underexplored. To our knowledge, this is the first study to directly link deficits in nurses’ knowledge and attitudes to reduced caring behaviors in this population. Currently, evidence indicates that training programs to improve nurses’ knowledge and attitudes— especially when paired with organizational and role-specific support—can significantly improve the quality of care nurses provide to people with substance use disorders (24, 32, 36). Given persistent gaps in hospital-based care for people with OUD, there is an urgent need for OUD-specific educational interventions to enhance nurses’ knowledge, and reshape their attitudes to foster more compassionate, evidence-based care for people with OUD (2, 18, 36).

In this study, we also examined whether nurses’ perceived knowledge, attitudes, and caring behaviors varied based on demographic and work-related factors. We identified significant differences in nurses’ knowledge and attitudes across several work-related variables, including work experience, employment status, and hospital type. Specifically, nurses employed in community hospitals reported lower knowledge and less favorable attitudes compared to those in teaching or university hospitals. Part-time nurses exhibited lower knowledge and more negative attitudes than their per diem counterparts. Interestingly, mid-career nurses (6–10 years of experience) demonstrated greater knowledge and more positive attitudes than those with longer careers. Regarding caring behaviors, nurses with only an RN license working at the bedside exhibited stronger caring behaviors than those in educational roles. These findings contribute to the existing literature by highlighting how workplace factors influence nurses’ preparedness to care for people with OUD. Previous studies have identified several factors affecting nurses’ knowledge and attitudes, such as stigma, compassion fatigue, lack of OUD-specific training, clinical exposure, demographic characteristics, institutional culture, and organizational support (1, 8, 11, 15, 17, 18, 26). While earlier research suggests that increased age and years of nursing experience improve attitudes and knowledge (2, 17, 18), our findings diverge, as these factors did not significantly impact attitudes or knowledge levels in our sample. A study of emergency department nurses found that younger, less experienced nurses reported more favorable attitudes and greater knowledge gains following training compared to their older, more experienced counterparts—highlighting the complex relationship between age, work experience, and learning responsiveness (30). These results underscore the multifactorial nature of workplace influences on nurses’ readiness to care for people with OUD. Factors such as clinical exposure frequency, institutional culture, and access to professional development may all contribute to variations in knowledge, attitudes, and caring behaviors (2, 26). Nurses in different healthcare settings, such as community versus university hospitals, are likely to face diverse policies, patient demographics, and training opportunities, all of which influence their practice (27). These complexities emphasize the need for continued research on how individual, interpersonal, organizational, and policy-level factors shape nurses’ attitudes, knowledge, and training needs for OUD care (2).

We would like to acknowledge limitations. First, due to the cross-sectional design of this study, causality cannot be established. Additionally, while well-validated instruments were used to measure the construct of knowledge, attitudes, and caring behaviors, these self-reported measures may be subject to response bias, with participants potentially overestimating or underestimating their knowledge and attitudes or caring behaviors. Moreover, using a non-probability sampling method limits the generalizability of the findings to the target population of inpatient nurses. Lastly, the study cohort was predominantly female (98%), which does not fully reflect the diversity of the current nursing workforce, potentially limiting the applicability of the results to a more representative nursing population. Future studies should consider a larger, more diverse, and representative sample to enhance the generalizability of the findings.

### Conclusions and Implications

In conclusion, deficits in nurses’ perceived knowledge and attitudes significantly influence their caring behaviors toward people with OUD. We found several workplace factors associated with variations in nurses’ knowledge, attitudes, and caring behaviors, but holding an RN license emerged as the only significant workplace factor contributing to the relationship between knowledge and attitude deficits and reduced caring. These findings have important implications for nursing practice and hospital-based OUD care. Given the ongoing challenges in caring for patients with OUD—compounded by healthcare stigma, compassion fatigue, knowledge deficits, and the lack of OUD-specific training for nurses (2, 5, 11, 16, 18, 28)—there is an urgent need to develop and implement a targeted and scalable empathy-based training program to address gaps in nursing OUD care (16, 26). This educational intervention should be user-friendly, seamlessly integrated into electronic medical record systems, and specifically designed to meet the demands of fast-paced nursing work environments. Such a program must emphasize evidence-based practice and stigma reduction and should be systematically implemented across all levels of nursing practice—from bedside clinicians to leadership positions. Over time, such interventions could help foster a culture of continuous learning, while enhancing nurses’ knowledge, shifting attitudes, and strengthening their caring behaviors toward people with OUD. Finally, with enhanced OUD-specific knowledge, nurses will be better positioned to advocate for early initiation of MOUD during intake or medication reconciliation, support coordinated team-based care, and promote structured discharge planning for patients with substance use disorders (4,16, 19). As the largest segment of the healthcare workforce, nurses are uniquely positioned to drive these improvements. Therefore, strategic investment in their education and support is essential for reshaping hospital-based OUD care and empowering nurses to deliver compassionate, coordinated, and evidence-based OUD care (15, 26, 30, 33).

## Acknowledgments

We would like to thank the researchers and community partners at Tufts Medical Center, as well as our research collaborators at UMass Lowell, Lowell, MA, and Boston Medical Center, Boston, MA. We also thank Merrimack College for providing funding and grant support for this research project.

## Supporting Information

### Data availability statement

All relevant data are within the manuscript and a link has been provided below to the study’s dataset.

### Link to the Data file

for Nursing Knowledge/Attitudes and Caring Behavior Study: https://www.openicpsr.org/openicpsr/workspace?goToPath=/openicpsr/230081&goToLevel=project

### Funding Sources

This work was supported by a Faculty Development Grant [START], awarded to Inyene Essien-Aleksi by Merrimack College in North Andover, Massachusetts.

### Author Contributions

Conceptualization: Inyene E. Essien-Aleksi, PhD, RN, Danielle Leone-Sheehan, PhD, RN, Yuan Zhang, PhD, RN.

Data curation: Inyene E. Essien-Aleksi, PhD, RN, Yuan Zhang, PhD, RN

Formal analysis: Inyene E. Essien-Aleksi, PhD, RN

Funding acquisition: Inyene E. Essien-Aleksi, PhD, RN.

Investigation: Inyene E. Essien-Aleksi, PhD, RN, Danielle Leone-Sheehan, PhD, RN, Yuan Zhang, PhD, RN, Tracie McPadden, DNP, Leslie Rideout, PhD, Michael Martin, MSN, Paula-Jo Beniers, MSN, Amy Lund, RN.

Methodology: Inyene E. Essien-Aleksi, PhD, RN, Danielle Leone-Sheehan, PhD, RN, Yuan Zhang, PhD, RN.

Project Administration: Inyene E. Essien-Aleksi, PhD, RN, Danielle Leone-Sheehan, PhD, RN, Leslie Rideout, PhD, Michael Martin, MSN, Paula-Jo Beniers, MSN, Amy Lund, RN, Yuan Zhang, PhD, RN

Software: SAS (version 9.4)

Supervision/mentor: Yuan Zhang, PhD, RN

Validation: Inyene E. Essien-Aleksi, PhD, RN, Danielle Leone-Sheehan, PhD, RN, Leslie Rideout, PhD, RN,Yuan Zhang, PhD, RN

Visualization: Inyene E. Essien-Aleksi, PhD, RN, Don Roosan, PhD, Yuan Zhang, PhD, RN, Alysse Wurcel, MD

Preparation: Inyene Essien-Aleksi, PhD, RN.

Writing-review editing: Inyene E. Essien-Aleksi, PhD, RN, Yuan Zhang, PhD, RN, Don Roosan, PhD, Danielle Leone-Sheehan, PhD, RN, Leslie Rideout, PhD, RN, Alysse Wurcel, MD

